# Demographic and socioeconomic determinants of access to care: A subgroup disparity analysis using new equity-focused measurements

**DOI:** 10.1101/2023.04.28.23289172

**Authors:** Miao Qi, Henrique Santos, Paulo Pinheiro, Deborah L. McGuinness, Kristin P. Bennett

## Abstract

Disparities in healthcare access and utilization associated with demographic and socioeconomic status hinder advancement of health equity. Thus, we designed a novel equity-focused approach to quantify variations of healthcare access/utilization from the expectation in national target populations. We additionally applied survey-weighted logistic regression models, to identify factors associated with usage of a particular type of health care. To facilitate generation of analysis datasets, we built an NHANES knowledge graph to help automate source-level dynamic analyses across different survey years and subjects’ characteristics.

We performed a cross-sectional subgroup disparity analysis of 2013-2018 National Health and Nutrition Examination Survey (NHANES) on U.S. adults for receipt of diabetes treatments and vaccines against Hepatitis A (HAV), Hepatitis B (HBV), and Human Papilloma (HPV). Results show that, in populations with hemoglobin A1c level *≥* 6%, patients with non-private insurance were less likely to receive newer and more beneficial antidiabetic medications; being Asian further exacerbated these disparities. For widely used drugs such as insulin, Asians experienced insignificant disparities in odds of prescription compared to White patients but received highly inadequate treatments with regard to their distribution in U.S. diabetic population. Vaccination rates were associated with some demographic/socioeconomic factors but not the others at different degrees for different diseases. For instance, equity scores increase with rising education levels for HBV while decrease with rising wealth levels for HPV. Among women vaccinated against HPV, minorities and poor communities usually received Cervarix while non-Hispanic White and higher-income groups received the more comprehensive Gardasil vaccine.

Our study identified and quantified the impact of determinants of healthcare utilization for antidiabetic medications and vaccinations. Our new methods for semantics-aware disparity analysis of NHANES data could be readily generalized to other public health goals to support more rapid identification of disparities and development of policies, thus advancing health equity.

## Introduction

Ensuring fair and equitable health care access and utilization for people with the same health needs, regardless of their demographic and socioeconomic status, has become a primary goal of the Centers for Disease Control and Prevention (CDC) to eliminate health inequity and improve the health system [1]. However, studies have consistently shown differences in health care access and utilization accounted for by demographic characteristics (e.g., race, ethnicity, and gender) and socioeconomic characteristics (e.g., insurance type, education degree, and poverty level) are important factors that may lead to dramatic life expectancy differences among subpopulations. For instance, the drug overdose mortality rate was 32.1% for individuals with Bachelor’s degree or more and 88.0% for those with lower education attainment in 2015-2019 [2]; diabetic patients from Black and Hispanic communities or lower income areas were reported having a lower chance to receive guideline-recommended treatments [2].

Despite considerable progress made, drug inaccessibility for some cohorts of the population with a specific disease, such as type 2 diabetes mellitus (T2DM), was still shown to be associated with poor health outcomes [3], leaving patients at risk for serious medical issues. Similarly, the COVID-19 pandemic has again exposed widened inequities in vaccine access that led to preventable deaths [4].

Essential medicines and vaccines are the most frequently used healthcare services, and they have proven to effectively treat, manage, and prevent many diseases [5]. So we explored utilization of antidiabetic medications and vaccines commonly recommended by CDC as examples to demonstrate the application of our approach to evaluating equity of access in healthcare.

To identify and evaluate potential significant inequities in health care utilization, we develop an approach to decide whether a subgroup received sufficient health care services compared to their share in the target population of subjects who need the services. Here, equity aims to allocate resources/opportunities accordingly across subgroups based on their different circumstances to achieve the same outcomes while equality means to provide the same resources/opportunities to different subgroups [6]. Prior work on equity of randomized clinical trials, developed equity metrics and significance tests that quantified if subgroups in a target population were observed to have disparate access to a clinical trial [7]. These equity metrics and methods can be immediately generalized to the problem of identifying if subgroups in a target population exhibited disparate access to a specified treatment. For example, we examine disparities in vaccine usage for subgroups defined by race/ethnicity and socioeconomic status with respect to the United States population. We select one or more demographic and socioeconomic covariates (referred to as sensitive covariates) and then examine subgroups defined by one or more of these attributes. The proposed equity-focused approach identifies subgroups defined by multiple covariates which are receiving less or different treatment than expected given the target population.

We observe that our novel equity-focused method adds insights beyond those found by logistic regression (LR) [8]. As commonly done in the health domain, we use LR to identify covariates associated with increased odds of specific healthcare utilization.

Specifically, we use LR to estimate if independent sensitive covariates have a significant relationship (quantified by odds ratios (ORs)) with the outcome treatment variable adjusted for confounding factors. LR uses no information about the target population requiring care and it typically doesn’t model interactions between the sensitive covariates. The proposed equity-focused approach identifies subgroups, defined using potentially multiple covariates, that receive disparate health care with respect to a specific target population.

We examined distributions of access to needed care, which include antidiabetic drugs and vaccines for the hepatitis A (HAV), the hepatitis B (HBV), and the human papillomavirus (HPV), across U.S. adults and studied associations with demographic features (e.g., gender and race/ethnicity) and socioeconomic determinants of health (e.g., education attainment, poverty income ratio (PIR), and insurance status) on patients’ reception of care.

For supporting these analysis requirements, we developed an approach based on Semantic Web technologies [9] to model and integrate NHANES data as a Knowledge Graph. This approach formalizes NHANES survey knowledge that is present in the original datasets and online documentation as published by the CDC into machine-interpretable semantic data dictionaries [10] (SDDs), including codebooks. The constructed SDDs and related data were then used in a novel semantics-aware data integration framework to build the NHANES Knowledge Graph and to allow the generation of prepared datasets according to user selection of variables of interest. This approach helps automate source-level data preparation for dynamic analysis across survey years and subjects’ characteristics for diverse research objectives. The semantically-supported equity analysis can facilitate future equity analysis of services beyond those in this study.

Our study identified and quantified potential demographic and socioeconomic determinants of health care utilization for a range of healthcare problems. For example, patients with non-private insurance might miss opportunities to be prescribed newer and more beneficial antidiabetic medications, and being non-Hispanic Asians further exacerbates disparities in types of prescribed antidiabetic drugs compared to other racial/ethnic groups; minorities seem to have more access to HAV and HBV vaccines while non-Hispanic White population got more HPV vaccines; specifically for HPV, minorities and poor communities usually got Cervarix while non-Hispanic White and higher-income populations were more likely to get Gardasil, which is a more comprehensive vaccine than Cervarix. The evidence suggests that analysis of equitable access and utilization of care should be performed routinely to support major public health goals and be considered in policies, thus advancing health equity and supporting better health for all.

Our main contributions include design and utilization of a health equity assessment methodology and infrastructure which:

- Introduces an approach for identifying subgroups with disparate access to health care services with respect to a target population.
- Determines/quantifies putative effects of various determinants of health on equity of access to health care using new analysis and visualization methods.
- Creates a novel semantics-aware data integration framework including the NHANES ontology for assisting automation of source-level dynamic analyses across surveys for different research objectives.
- Applies the health equity assessment framework on diverse health problems by showing observed differences in health care access for both vaccines and type-2 diabetes medications.

In this paper, we first introduce the approaches that measure disparities of health care utilization between groups, including the novel subgroup disparity approach built upon our health equity framework [7] and the traditional logistic regression model.

Then, we discuss details on selected demographic and socioeconomic determinants of health and on the method to estimate study and target populations from NHANES. Next, we applied the approaches to evaluate utilization of antidiabetic drugs, HAV, HBV, and HPV vaccines. Finally, we discuss the advantages and limitations of our equity-based approach and point out some potential directions of future work.

## Materials and methods

To identify the underlying determinants of inequitable health care access/utilization and to monitor health disparities in medication and vaccine utilization, we examined distributions of utilization of needed care, which include antidiabetic drugs, HAV, HBV, and HPV, across U.S. adults. We developed an approach based on the equity framework from our previous work [7] to study determined effects of demographic features (e.g., gender and race/ethnicity) and socioeconomic determinants of health (e.g. education attainment, poverty income ratio (PIR), and insurance status) on patients’ utilization of care in addition to logistic regression.

For medications, the outcome variable of interest is whether the participant was prescribed a drug from a specific Multum drug/ingredient therapeutic category [11]. For vaccines, the outcome variable of interest is whether subjects received vaccine. Type of HPV vaccine received is also evaluated for HPV but constrained to people who received HPV vaccine.

### Subgroup disparity analysis

Our analyses on equitable health care utilization are based on two approaches: a new equity metric-based method and a logistic regression model frequently used for determinants of health.

### Equity metric-based novel approach

Our equity metric-based approach is applied to decide whether the share of healthcare access and utilization across subgroups differentiated by demographic and socioeconomic factors of interest were proportionate to their share of the target population. To get clinically significant findings, we use subpopulation analysis to control for patient characteristics that differentiate drug prescribing of physicians by following treatment guidelines. For example, since HbA1c level is clinically associated with the decision on prescribed antidiabetic drug class, we analyzed the effect of demographic and socioeconomic factors within subpopulations that have similar HbA1c levels.

Multivariable conditions, such as both HbA1c level and the Charlson Comorbidity Index (CCI) level [12], can be applied to obtain a subpopulation-level equity heatmap for each subgroup defined over the conditioned attributes. This method quantifies disparities, providing opportunities to monitor and improve health equity improvement.

In this approach, a statistical analysis is performed to test if the disparities between the subgroups’ observed and target utilization rates of health care services are significant. If the p-value is smaller than the significance threshold 0.05, then we define that the subgroup received adequate service they needed; if *p − value ≥* 0.05, then an equity metric [7] is applied to calculate a score that determines whether the utilization rate of health care service is inappropriate for the population. For each equity metric, an upper threshold *τ_u_* and a lower threshold *τ_l_* of metric values are defined to categorize metric scores into different equity levels. In our evaluation, the Log Disparity metric [7],

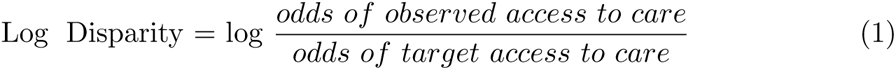

is used. The threshold is *τ*_log disparity_ = *−log*(1 *− τ_rule_*), where *τ_rule_ ∈* [0, 1]. By following the 80% rule [13], our *τ_l_* = *−log*(0.8). The *tau_u_* is user-defined and was selected to be *−log*(0.6) in our experiment.

The color description of equity evaluation heatmaps is available in Table 1. The severity of inequity is mapped to color to help us visualize where disadvantaged subpopulations are, discover social determinants of health care utilization, or identify communities that need immediate intervention.

**Table 1.** Color description for heatmap.

| Color | Description | Equity Score |
| --- | --- | --- |
| Red | Absent | * |
| Orange | Highly Inadequate | $< -\tau_u$ |
| Light orange | Inadequate | $[-\tau_u, -\tau_l)$ |
| Teal | Adequate | $[-\tau_l, \tau_l)$ or $p > 0.05$ |
| Light blue | Abundant | $[\tau_l, \tau_u)$ |
| Blue | Highly Abundant | $\geq \tau_u$ |

### Multivariate logistic regression model

We also used the multivariate logistic regression model, which is a popular and widely used association analysis method in health domain [14–19], to determine effects of demographic features and socioeconomic determinants of health on the access and utilization of health care.

We construct the model as a function of race/ethnicity, age, gender, educational attainment, insurance type, poverty level, comorbidity severity based on CCI, and HbA1c condition. The OR is the odds of drug/vaccination access and utilization of a subgroup divided by the odds of the same healthcare source access and utilization in a reference group. Reference groups are non-Hispanic White, male, and private insurance for unordered categorical variables; reference groups are the lowest level of ordered ones (e.g., CCI=0, HbA1c condition *<* 6%). This method controls for the influence of approved time for usage by FDA across medications through age adjustment.

To apply logistic regression, we use the R package “svydiags” [20] to check the assumptions considering the complex survey design of NHANES. The assumptions of logistic regression are satisfied except that extreme outliers exist in our data. One limitation of logistic regression is that these assumptions are not always satisfied.

### Data source and study population

In the study, we analyzed the National Health and Nutrition Examination Survey (NHANES) demographic, socioeconomic, diabetes, and vaccination data for the 2013-2018 survey cycles [21]. All the NHANES programs were approved by the National Center for Health Statistics (NCHS) Ethics Review Board and the informed consent was signed by all subjects. The NHANES data used in the manuscript are de-identified and remain anonymous during the analysis. Therefore, no ethical approval for this secondary research was required. By using appropriate weighting through R survey package [22], a nationally representative population was estimated from NHANES surveys conducted and approved by the NCHS. All the The selection and use of weights depend on survey cycles and subjects’ characteristics used in the analysis.

When we first analyzed NHANES, we used the traditional approach of having scientists conduct the process of inspecting the data, reading the data dictionaries, and confronting their interpretation of the data against online documentation and the data understanding developed by other scientists. By contrasting one interpretation against other interpretations, we collectively acquired a clarification on the exact meaning of the raw data that is often lost after similar analyses are done. This time, however, we have preserved this data understanding by translating human-level data dictionaries and codebooks into machine-level semantic data dictionaries and codebooks. Further, we have used these machine-level documents along with raw data and a novel semantic data integration infrastructure [23] to build an NHANES knowledge graph [24] that is publicly available at http://nhanes.eci.ufmg.br:9000/hadatac.

With the knowledge graph and infrastructure in place, we have repeated our original analysis using the traditional approach and used it to compare against the results produced from datasets automatically generated (prepared) from the infrastructure. The obtained results were identical, indicating that future analysis of NHANES data can have their expensive data preparation work expedited through the use of our semantic infrastructure.

### Semantic-aware data integration approach

In NHANES, we experience common disconnects between data and knowledge that had to be addressed. For instance, data dictionaries (DDs) used to support data understanding by humans cannot be easily leveraged by machines. The DDs include natural language descriptions of the variables composing a dataset, as well as codebooks for select variables where codes are used instead of direct values. From the DDs we have created Semantic Data Dictionaries (SDDs) [10] where an identified associated entity, attributes, unit, set of code book values, time, space, and provenance properties are used to formalize the knowledge related to each variable.

During the creation of SDDs, we further used ontology engineering [25] best practices to build an NHANES ontology^1^. Our ontology references appropriate terminology from other well-cited ontologies, adds NHANES terms that are not well described in existing ontologies, and is also used to integrate code book terms to existing databases such as ICD10-CM and RXNORM. For each codebook value, an OWL class is defined with the appropriate metadata (e.g., description, source variable, code value).

Our semantic enhancements have the potential to affect data interpretation. For example, some of the codebooks, when human-interpreted, provide categorical values that group a range of values into a single code. One example is the “education level” variables which have values that include “less than 9th grade” and “9-11th grade” for survey participants, and “less than high school degree” for household reference persons. In this case, the more discrete ranges in the survey participants’ codebook were defined as subclasses of the broader definition in the household reference persons’ codebook.

Fig 1 shows part of our ontology covering some of the education classes mentioned above. This principle was applied to other variables where codebooks could be interpreted to identify similar relationships, and the results were incorporated into our NHANES ontology.

**Fig 1.**
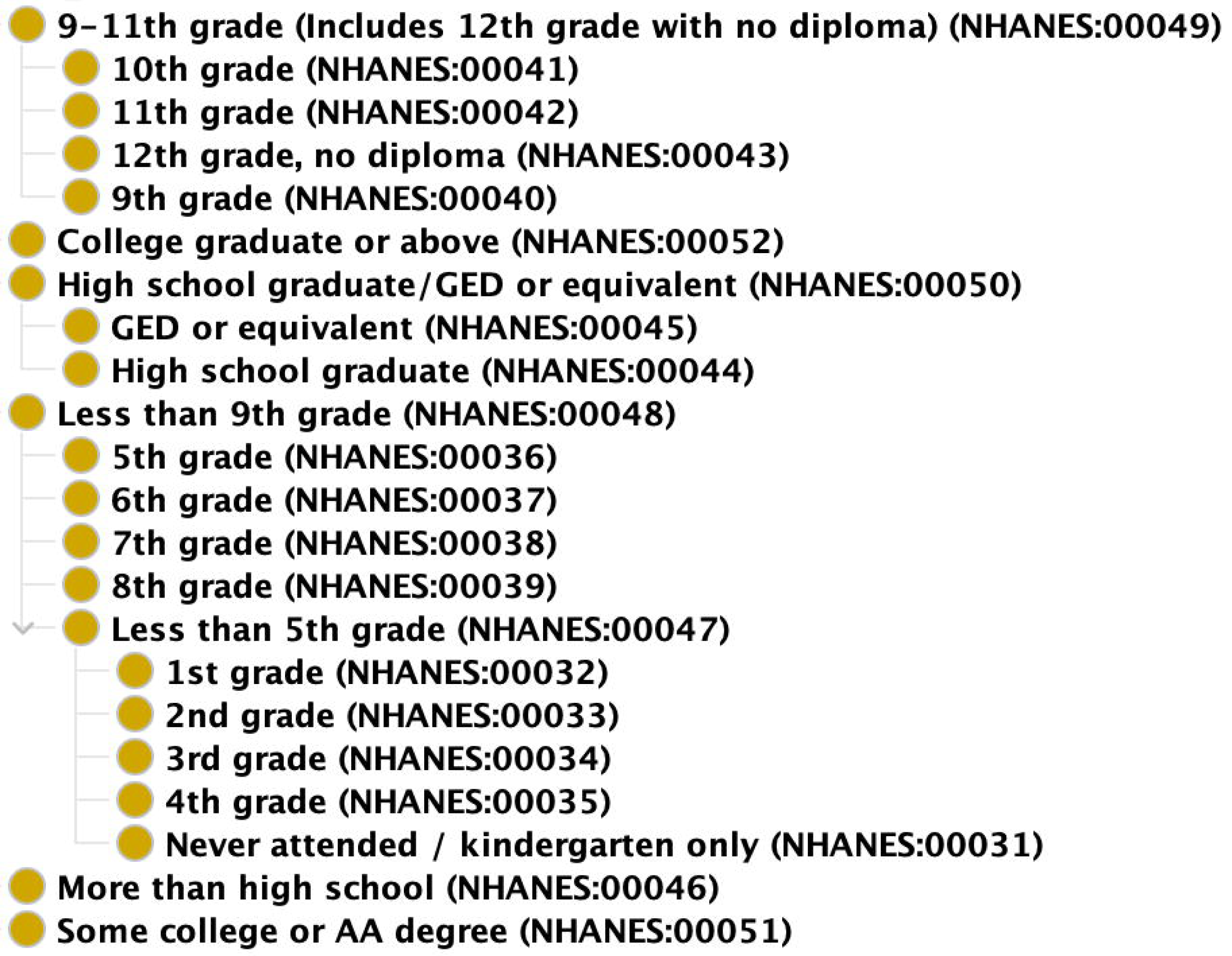
NHANES Ontology. Part of our NHANES Ontology showcasing the harmonization of the several education levels across datasets.

### Vaccination

We use the three types of vaccines (i.e., HAV, HBV, and HPV) available in NHANES immunization documents to identify and evaluate potential inequitable access and utilization of immunization services experienced by some subgroups.

The demographic characteristics used in our analysis include self-reported age, gender (male, female), and race/ethnicity (non-Hispanic White, non-Hispanic Black, non-Hispanic Asian, Hispanic, or other/unknown). The socioeconomic characteristics include education level, insurance type, and the ratio of family income to poverty. The categorization of these variables follows. Education levels available from NHANES include less than 9th grade, 9-11th grade (includes 12th grade with no diploma), high school graduate/GED or equivalent, some college or AA degree, and college graduate or above. Health insurance status was classified as private insurance, Medicare, Medicaid, and other non-private insurance. The ratio of family income to poverty were categorized into poor (*<* 1), near poor (1-1.9), middle income (2-3.9), and higher income (*≥* 4).

Data preparation issues are also present in these socioeconomic determinants. For instance, the Insurance datasets contain several variables used to fully characterize insurance coverage. Each variable contains the participation status of the survey participant in one specific type of insurance (such as Medicaid, Medicare, Private insurance, etc.). However, we understand insurance coverage as not the value of a single variable but the combination of several variables insurance-related variables, all contributing to the insurance coverage attribute of the participant. For example, we can only infer if a person does not have insurance coverage if all variables contain the information of not being covered. With the use of our NHANES semantic infrastructure, insurance coverage is available as a multivalued variable.

For HAV/HBV vaccination, we included subjects over age 20 and not pregnant based on the urine pregnancy test result; for HPV vaccination, only participants aged between 20 and 59 were analyzed due to the NHANES design [26].

### Antidiabetic medication

We use antidiabetic medications as an example to identify and evaluate potential over-/under-prescribing or over-/under-use of certain types of medications to treat chronic conditions experienced by some populations.

The same demographic (i.e., age, gender, and race/ethnicity) and socioeconomic (i.e., education level, insurance type, and ratio of family income to poverty) factors for vaccinations are used for T2DM. However, the age categorization is updated to 20-45, 46-64, *≥* 65 [27] according to disease domain knowledge from CDC.

For antidiabetic drug analysis, we included subjects with known T2DM, aged over 20, and nonpregnant. Additionally, we focused on T2DM treatments by excluding medications used for type 1 diabetes and for prevention. It was important for the semantic infrastructure to differentiate between the use of drugs to treat disease and to prevent disease, which is convoluted in the raw data since this is a distinction that was introduced later in the study. In our modeling, drug usages for treatment and prevention are separate variables.

To explore the effect of demographic features and socioeconomic determinants of health on patients’ access to American Diabetes Association (ADA) recommended T2DM treatments [28], we included 60 antidiabetic drugs available in NHANES and grouped them based on the nested 3-level Multum Lexicon therapeutic classification scheme [11]. The 10 first level categories we used include meglitinides, sodium-glucose cotransporter type 2 inhibitors (SGLT2is), sulfonylureas (SUs), biguanides, dipeptidyl peptidase IV inhibitors (DPP-4is), insulin, thiazolidinediones (TZDs), glucagon like peptide-1 receptor agonists (GLP-1RAs), *α*-glucosidase inhibitors (AGIs), and antidiabetic combinations. The second level categorization includes details of antidiabetic combinations, and the third level categorization further categorized the drugs based on ingredients.

According to the 2022 ADA guideline [28], treatment recommendation for adults with T2DM depends on comorbidities. So, 15 common comorbidities were considered in our analysis: hypertension, asthma, arthritis, gout, congestive heart failure, coronary heart disease, heart attack, stroke, emphysema, chronic bronchitis, cancer, liver disease, COPD (chronic obstructive pulmonary disease), kidney disease, and diabetic retinopathy. The CCI scores [12] of participants were calculated as an indicator of severity of comorbidity and were categorized into four levels: none (CCI score = 0); mild (CCI scores of 1–2); moderate (CCI scores of 3–4); and severe (with CCI scores *≥* 5) [29]. Another factor that influences antidiabetic medication prescription is Hemoglobin A1C (HbA1c), which was categorized into *<* 6%, 6% - 7%, 7% - 9%, and *≥* 9% [30].

## Results

By comparing both reference groups and target populations, the findings suggest that different determinants of healthcare access and utilization exist regarding resources/services of different health needs.

### Impact of demographic and socioeconomic factors on vaccination utilization

Table 2 displays results from our logistic regression model of vaccination coverage for racial/ethnic subgroups (reference: non-Hispanic White), for different income subgroups (reference: poor population), for subgroups of different insurance types (reference: private insurance), and for subgroups with different educational attainments (reference: less than 9th grade). Minorities were more likely to be vaccinated against HAV/HBV while no vaccination coverage disparity was observed for HPV. For example, non-Hispanic Asian subjects experience an increase of 17% in the odds of getting HAV vaccine and an increase of 10% in the odds of getting HBV compared to non-Hispanic White subjects. Similar analyses were performed on other factors of interest but with different levels of disparities. For instance, the higher-income population experienced an increase of 4% in the odds for HAV vaccination; adults with an education level of 9-11th grade have an increase of 8% for HAV and 14% for HBV vaccinations compared to the subgroup of lower education level.

**Table 2.** Associations between population groups and vaccination by race/ethnicity, poverty level, insurance type, and education level in the U.S.

| Vaccine | HAV | HBV | HPV |
| --- | --- | --- | --- |
| NH Black | 1.09*(1.03, 1.16) | 1.05 (1.00, 1.10) | 0.99 (0.94, 1.04) |
| NH Asian | 1.17*(1.08, 1.26) | 1.10*(1.04, 1.17) | 0.94*(0.89, 0.99) |
| Hispanic | 1.13*(1.08,1.18) | 1.03 (0.97, 1.08) | 0.99 (0.94, 1.05) |
| Near poor | 1.01 (0.95, 1.06) | 1.00 (0.94, 1.06) | 1.02 (0.95, 1.08) |
| Middle income | 0.99 (0.95, 1.04) | 0.99 (0.94, 1.04) | 0.98 (0.94, 1.02) |
| Higher income | 1.04*(1.01, 1.08) | 1.03 (0.99, 1.07) | 0.98 (0.93, 1.04) |
| Medicaid | 1.09*(1.01, 1.17) | 1.03 (0.95, 1.12) | 1.02 (0.93, 1.11) |
| Medicare | 1.01 (0.94, 1.09) | 0.99 (0.90, 1.09) | 1.06 (0.93, 1.22) |
| Other insurance | 1.01 (0.92, 1.10) | 1.03 (0.93, 1.14) | 1.02 (0.96, 1.09) |
| 9-11th grade | 1.08*(1.02, 1.15) | 1.14*(1.06, 1.23) | 1.02 (0.96, 1.08) |
| High school graduate | 1.06 (1.00, 1.12) | 1.04 (0.97, 1.10) | 1.01 (0.96, 1.07) |
| Some college or AA degree | 1.01 (0.96, 1.05) | 0.98 (0.94, 1.03) | 1.00 (0.95, 1.06) |
| College graduate or above | 0.99 (0.94, 1.05) | 0.98 (0.94, 1.02) | 0.97 (0.92, 1.01) |

Reference groups: race/ethnicity = non-Hispanic White; poverty level = poor; insurance type = private insurance; education level = less than 9th grade. \* indicates statistically significant disparity with $p \leq 0.05$ .

Equity-focused results for the same independent and outcome variables as for the logistic regression are shown as heatmaps. In general, the trends are the same as those observed in Table 2 but it further detects the potential inequities in the reference population used for the logistic regression. Fig 2 shows that Hispanic population received adequate vaccination for all three types; non-Hispanic Asian population got more HAV and HBV vaccinations but less for HPV; no-Hispanic Black population received more HAV vaccination; and non-Hispanic White population did not get sufficient HAV vaccination. Furthermore, Fig 2D states that minorities tend to receive Cervarix more frequently than Gardasil vaccine which was designed to treat more types of infections.

**Fig 2.**
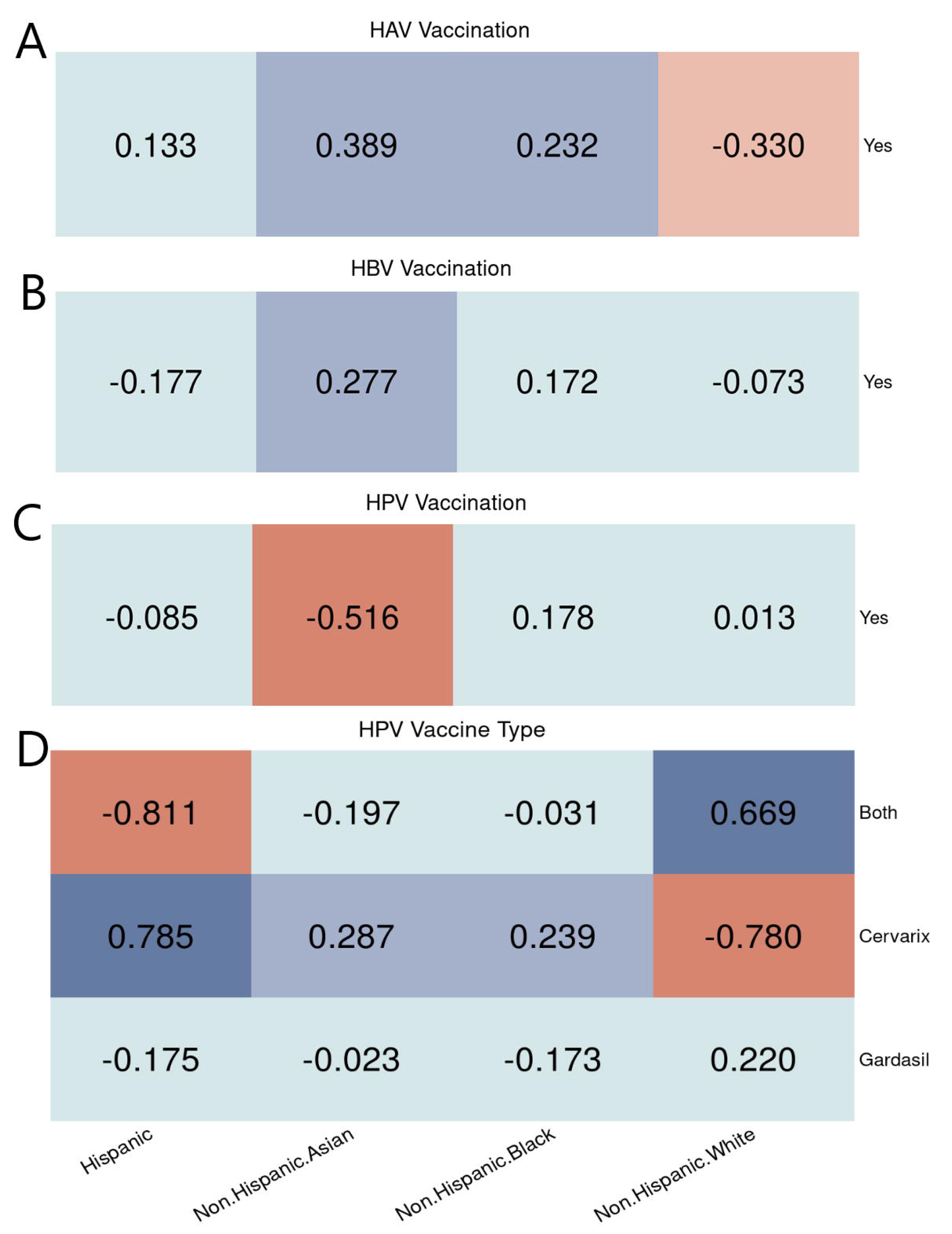
Racial/Ethnic equity evaluation on vaccination. Equity evaluation heatmaps of racial/ethnic disparities on different cases. A: HAV vaccination. B: HBV vaccination. C: HPV vaccination. D: HPV vaccine types used among people who got the vaccine.

The equity results found no significant relationships between poverty level and HAV or HBV vaccinations. For HPV, Fig 3 shows that poor people were more likely to receive HPV while the higher income population did not achieve the target rate for HPV as they needed; people with higher income were more likely to get Gardasil while the other groups got more Cervarix. For logistic regression, the higher income subgroup had increased adjusted odds of receiving HAV than the poor subgroup.

**Fig 3.**
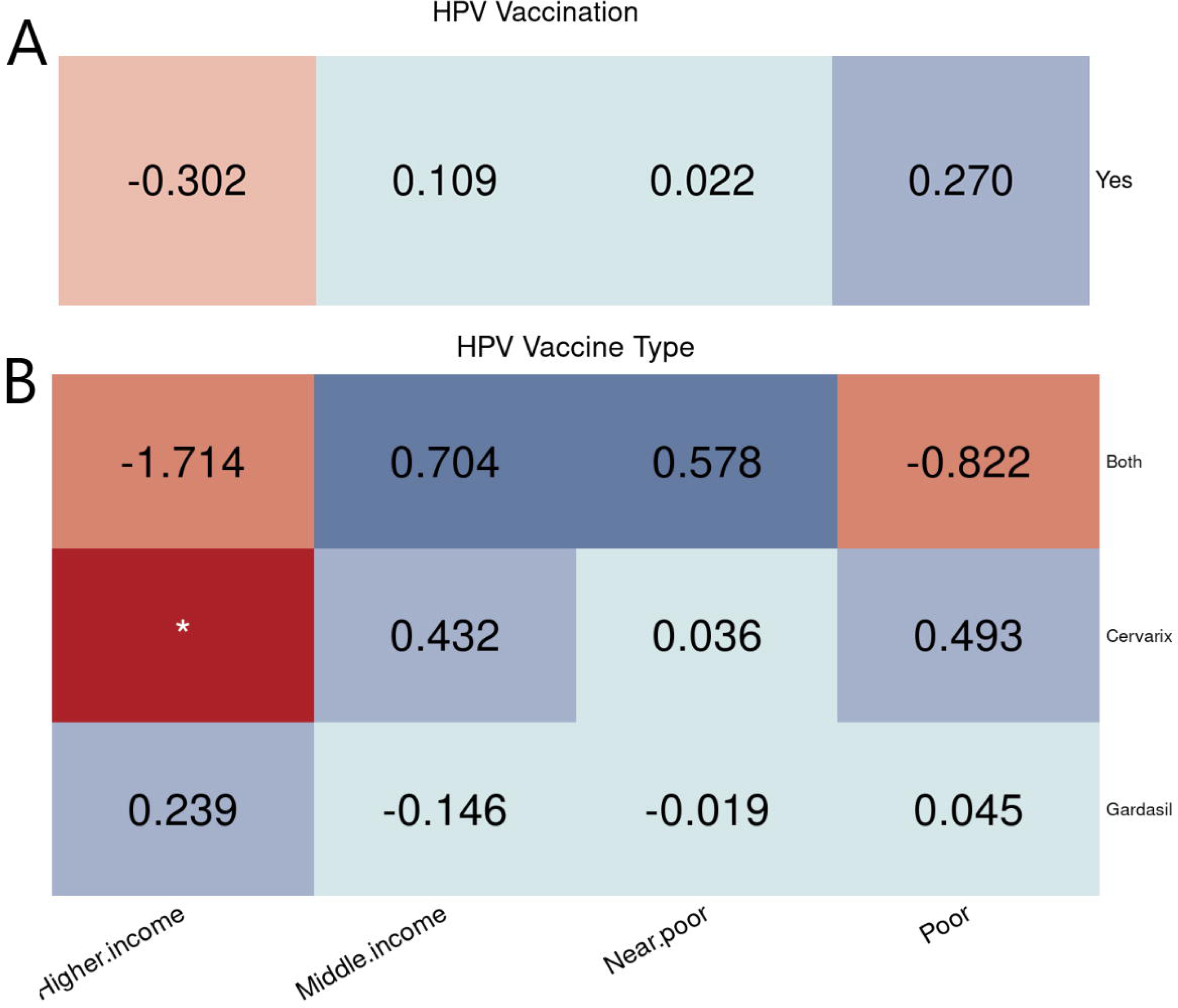
Poverty level equity evaluation on vaccination. Equity evaluation heatmaps of disparities by poverty level on different cases. A: HPV vaccination B: HPV vaccine types used among people who got the vaccine.

Analyses based on other demographic and socioeconomic factors were performed. For instance, as shown in Fig 4, regarding to education level, people who did not finish 9th grade tend to receive fewer vaccinations while people with higher education attainment got sufficient and sometimes more share of vaccinations.

**Fig 4.**
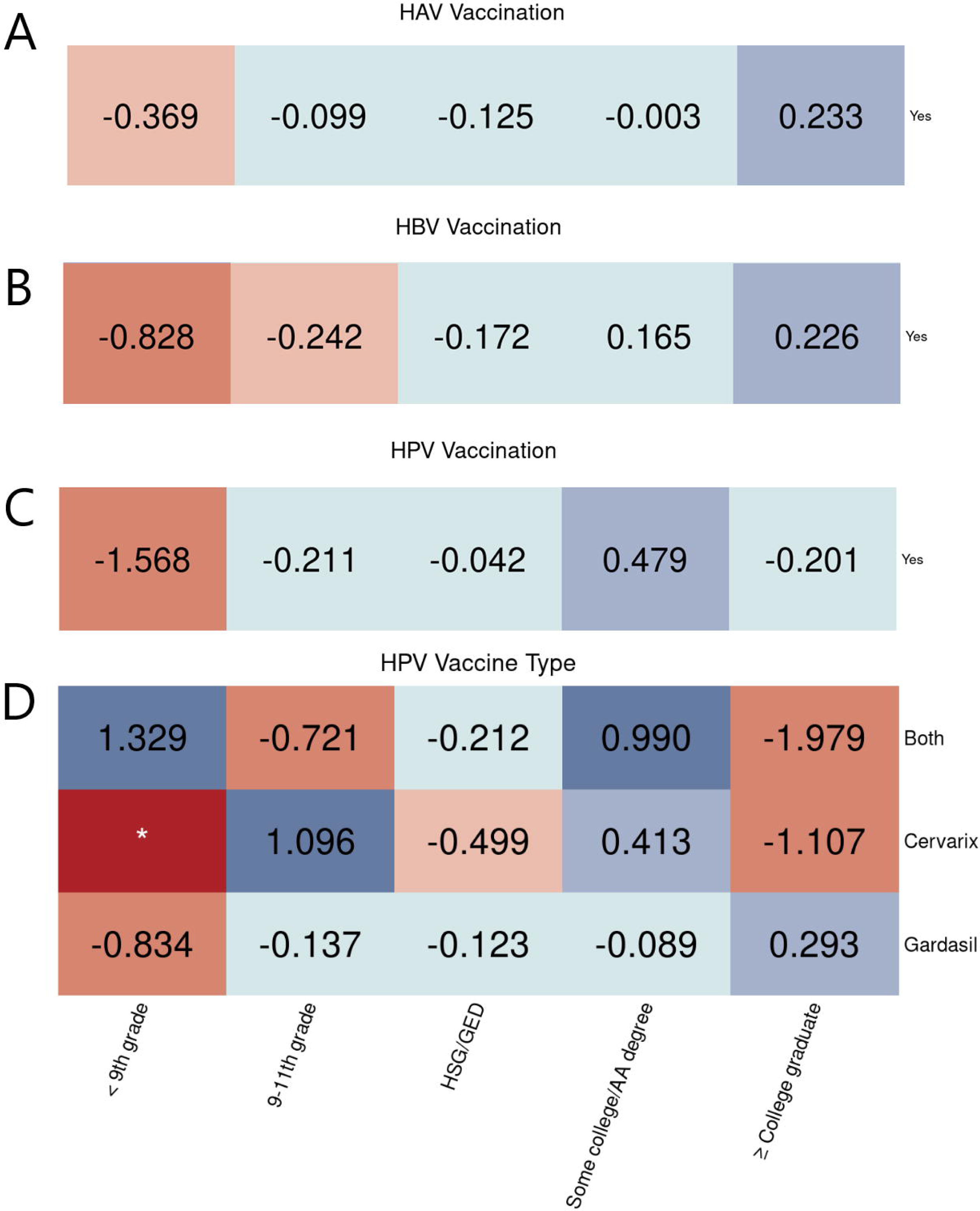
Education level equity evaluation on vaccination. Equity evaluation heatmaps of disparities by education level on different cases. A: HAV vaccination. B: HBV vaccination. C: HPV vaccination. D: HPV vaccine types used among people who got the vaccine.

### Impact of demographic and socioeconomic factors on antidiabetic drug utilization

Tables 3, 4, 5, and 6 display findings from the logistic regression model on antidiabetic medication utilization for the same subgroups as described for vaccines. For example, for the widely used drug such as biguanides, non-Hispanic Black patients had a 10% decrease in the odds of prescribing rate compared to the reference; for SGLT-2is, near poor population had a 2% increase in the odds of prescribing rate compare to the poor population; for all medication classes, no significant disparities were discovered between the utilization and insurance types by the logistic model. Some education-level-based disparities also existed, such as a 9% decrease in the odds of prescribing rate on biguanides for the population with college degree and higher education compared to those with less than a 9th grade education.

**Table 3.** Associations between population groups and diabetes drug use by race/ethnicity in the U.S.

| Medication Class | NH Black | NH Asian | Hispanic |
| --- | --- | --- | --- |
| AGIs | 1.00 (1.00,1.01) | 1.00 (0.99,1.00) | 1.00 (0.99,1.01) |
| Biguanides | 0.90*(0.81,0.99) | 1.02 (0.89,1.17) | 1.00 (0.91,1.11) |
| DPP-4is | 0.97 (0.92,1.03) | 1.04 (0.96,1.13) | 1.02 (0.95,1.10) |
| GLP-1RAs | 0.99 (0.94,1.04) | 0.94*(0.88,1.00) | 0.97 (0.93,1.01) |
| Insulin | 1.03 (0.91,1.16) | 0.89 (0.79,1.01) | 0.95 (0.87,1.05) |
| Meglitinides | 1.00 (0.99,1.02) | 1.00 (0.99,1.02) | 1.00 (0.99,1.01) |
| SGLT-2is | 1.02 (0.98,1.06) | 1.01 (0.96,1.06) | 1.00 (0.97,1.03) |
| SUs | 1.01 (0.91,1.11) | 1.14*(1.02,1.27) | 0.98 (0.89,1.08) |
| TZDs | 1.01 (0.97,1.06) | 0.96*(0.93,1.00) | 1.02 (0.98,1.06) |
| Combinations | 1.02 (0.97,1.07) | 1.01 (0.95,1.06) | 1.02 (0.97,1.08) |
| None | 0.99 (0.94,1.05) | 1.00 (0.95,1.05) | 1.03 (0.95,1.11) |

Reference group: non-Hispanic White. NH = non-Hispanic. \* indicates statistically significant disparity with $p \leq 0.05$ .

**Table 4.**
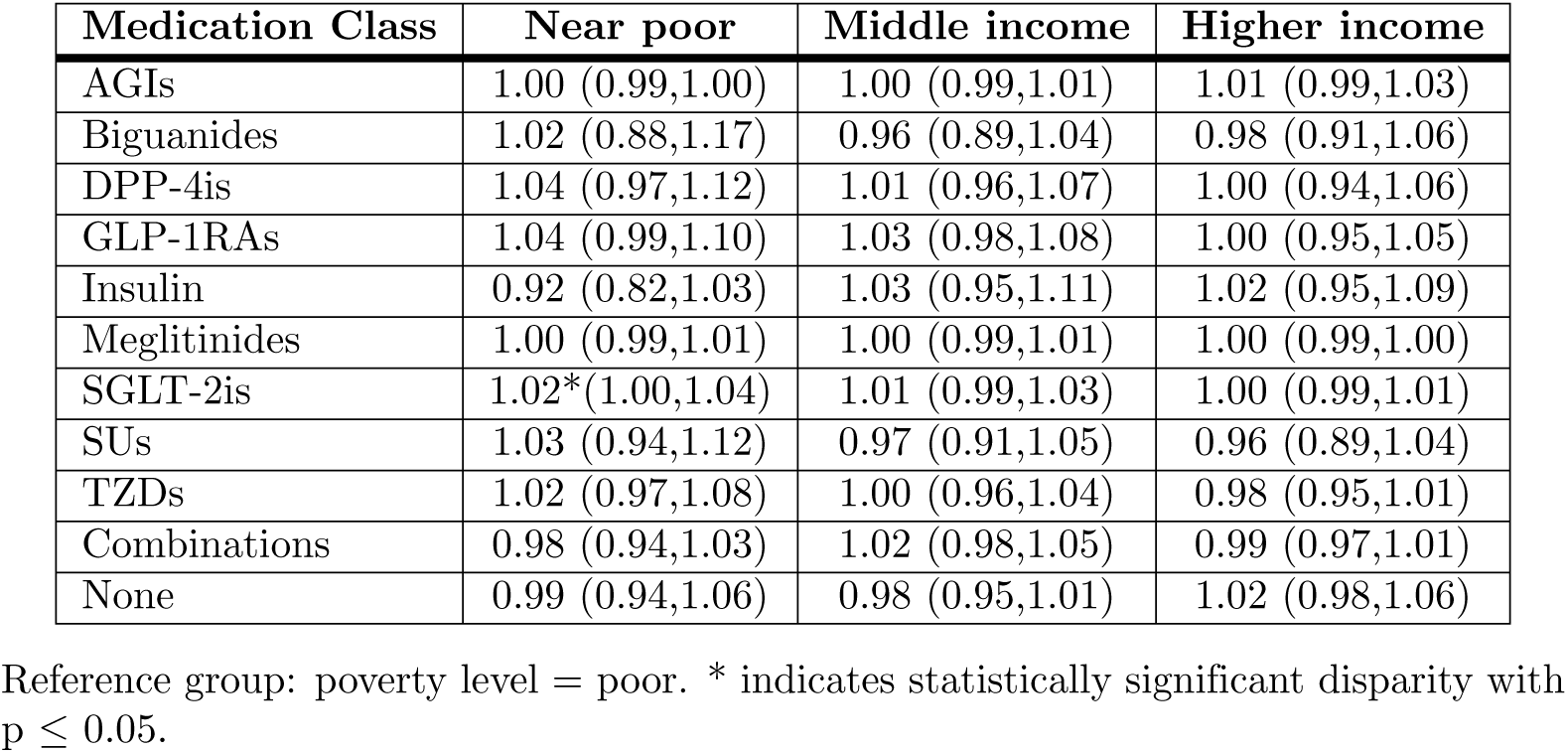
Associations between population groups and diabetes drug use by poverty level in the U.S.

| Medication Class | Near poor | Middle income | Higher income |
| --- | --- | --- | --- |
| AGIs | 1.00 (0.99,1.00) | 1.00 (0.99,1.01) | 1.01 (0.99,1.03) |
| Biguanides | 1.02 (0.88,1.17) | 0.96 (0.89,1.04) | 0.98 (0.91,1.06) |
| DPP-4is | 1.04 (0.97,1.12) | 1.01 (0.96,1.07) | 1.00 (0.94,1.06) |
| GLP-1RAs | 1.04 (0.99,1.10) | 1.03 (0.98,1.08) | 1.00 (0.95,1.05) |
| Insulin | 0.92 (0.82,1.03) | 1.03 (0.95,1.11) | 1.02 (0.95,1.09) |
| Meglitinides | 1.00 (0.99,1.01) | 1.00 (0.99,1.01) | 1.00 (0.99,1.00) |
| SGLT-2is | 1.02*(1.00,1.04) | 1.01 (0.99,1.03) | 1.00 (0.99,1.01) |
| SUs | 1.03 (0.94,1.12) | 0.97 (0.91,1.05) | 0.96 (0.89,1.04) |
| TZDs | 1.02 (0.97,1.08) | 1.00 (0.96,1.04) | 0.98 (0.95,1.01) |
| Combinations | 0.98 (0.94,1.03) | 1.02 (0.98,1.05) | 0.99 (0.97,1.01) |
| None | 0.99 (0.94,1.06) | 0.98 (0.95,1.01) | 1.02 (0.98,1.06) |

Reference group: poverty level = poor. \* indicates statistically significant disparity with $p \leq 0.05$ .

**Table 5.** Associations between population groups and diabetes drug use by insurance type in the U.S.

| Medication Class | Medicaid | Medicare | Other insurance |
| --- | --- | --- | --- |
| AGIs | 1.01 (0.99,1.02) | 1.00 (0.99,1.02) | 0.99 (0.98,1.01) |
| Biguanides | 0.94 (0.79,1.12) | 0.91 (0.79,1.06) | 0.99 (0.85,1.14) |
| DPP-4is | 0.99 (0.93,1.06) | 1.02 (0.95,1.08) | 1.02 (0.92,1.14) |
| GLP-1RAs | 1.01 (0.89,1.13) | 1.00 (0.93,1.09) | 0.95 (0.90,1.00) |
| Insulin | 1.07 (0.93,1.23) | 1.06 (0.96,1.17) | 0.96 (0.81,1.14) |
| Meglitinides | 1.01 (0.99,1.02) | 1.00 (0.99,1.01) | 1.00 (1.00,1.00) |
| SGLT-2is | 1.01 (0.97,1.05) | 1.00 (0.98,1.01) | 0.99 (0.95,1.03) |
| SUs | 0.94 (0.81,1.10) | 1.01 (0.92,1.10) | 0.91 (0.79,1.05) |
| TZDs | 0.98 (0.95,1.01) | 1.00 (0.95,1.05) | 0.98 (0.95,1.02) |
| Combinations | 1.01 (0.95,1.07) | 1.02 (0.98,1.08) | 0.99 (0.95,1.04) |
| None | 1.04 (0.94,1.14) | 1.01 (0.94,1.09) | 1.01 (0.91,1.11) |

Reference group: insurance type = private insurance. \* indicates statistically significant disparity with $p \leq 0.05$ .

**Table 6.** Associations between population groups and diabetes drug use by education level in the U.S. * indicates statistically significant disparity with p *≤* 0.05.

| Medication Class | 9-11th grade | High school graduate | Some college / AA degree | College graduate or above |
| --- | --- | --- | --- | --- |
| AGIs | 1.00(0.98,1.01) | 1.00(0.99,1.01) | 1.00(0.99,1.01) | 0.99(0.98,1.01) |
| Biguanides | 1.07(0.93,1.22) | 1.06(0.96,1.17) | 1.03(0.94,1.13) | 0.91*(0.82,1.00) |
| DPP-4is | 0.97(0.89,1.06) | 1.01(0.96,1.08) | 1.02(0.96,1.08) | 1.00(0.92,1.08) |
| GLP-1RAs | 1.04(0.98,1.10) | 1.06*(1.00,1.12) | 1.03(0.96,1.09) | 1.02(0.98,1.07) |
| Insulin | 1.02(0.94,1.11) | 0.98(0.90,1.07) | 1.03(0.96,1.11) | 1.02(0.96,1.10) |
| Meglitinides | 0.99(0.98,1.00) | 1.00(0.99,1.01) | 1.00(0.99,1.01) | 1.00(0.99,1.01) |
| SGLT-2is | 0.99(0.96,1.02) | 1.03(0.99,1.07) | 1.00(0.97,1.03) | 1.00(0.98,1.02) |
| SUs | 0.94(0.85,1.04) | 0.92(0.85,1.00) | 1.11*(1.03,1.18) | 1.03(0.94,1.13) |
| TZDs | 0.98(0.91,1.06) | 1.00(0.95,1.06) | 1.03(0.99,1.08) | 1.01(0.95,1.07) |
| Combinations | 1.01(0.95,1.07) | 1.03(0.98,1.09) | 0.99(0.96,1.03) | 1.01(0.98,1.04) |
| None | 0.99(0.95,1.03) | 0.98(0.94,1.03) | 0.98(0.95,1.01) | 1.04(1.00,1.09) |

Reference group: education level = less than 9th grade. \* indicates statistically significant disparity with $p \leq 0.05$ .

Findings based on the log disparity metric are displayed in Fig 5. Here, we applied the second-level Multum Lexicon therapeutic classification on antidiabetic medications for equity analysis. For instance, GLP-1RAs seem to be highly overprescribed to non-Hispanic Black patients and insulins may be highly underprescribed to non-Hispanic Asian patients.

**Fig 5.**
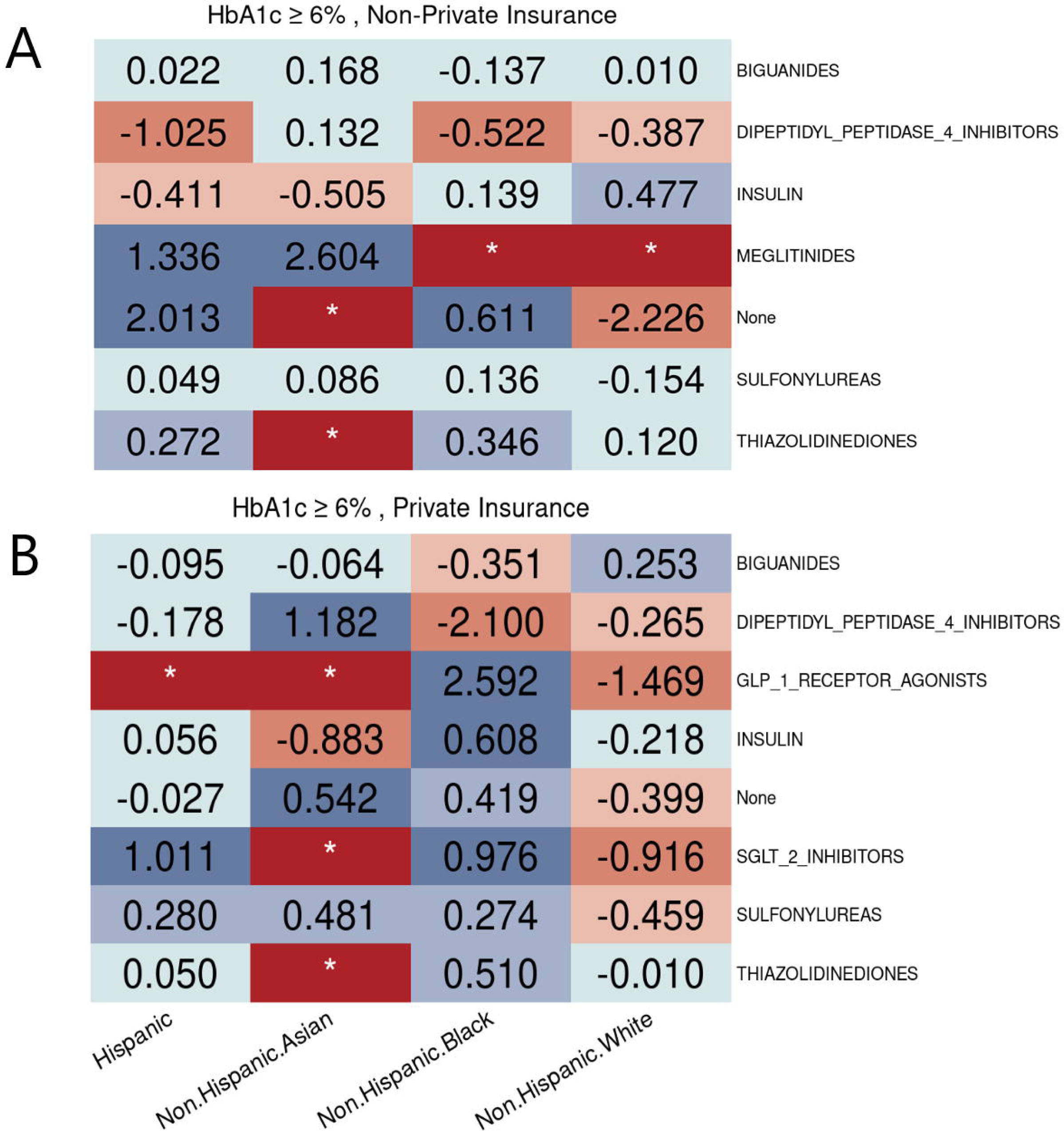
**Disparities of hyperglycemic medication utilization by race/ethnicity.**

From Fig 5, disparity is only observed in non-Hispanic Black patients for biguanides. When we further explored subpopulations with HbA1c *≥* 6% in Fig 6, we found that the disparities might be due to insurance types, where non-Hispanic Black patients with private insurance were less likely to be prescribed biguanides. This indicates that some factors only have effects on specific subgroups conditioned on disease-specific determinants of prescribing decisions. As shown in Fig 6, private insurance patients had access to newer antidiabetic drugs (i.e., SGLT2is and GLP-1RAs, which are the two new recommended treatments by ADA guideline 2022 [28]) while their counterparts with non-private insurance lost opportunities to get new effective treatments.

**Fig 6.**
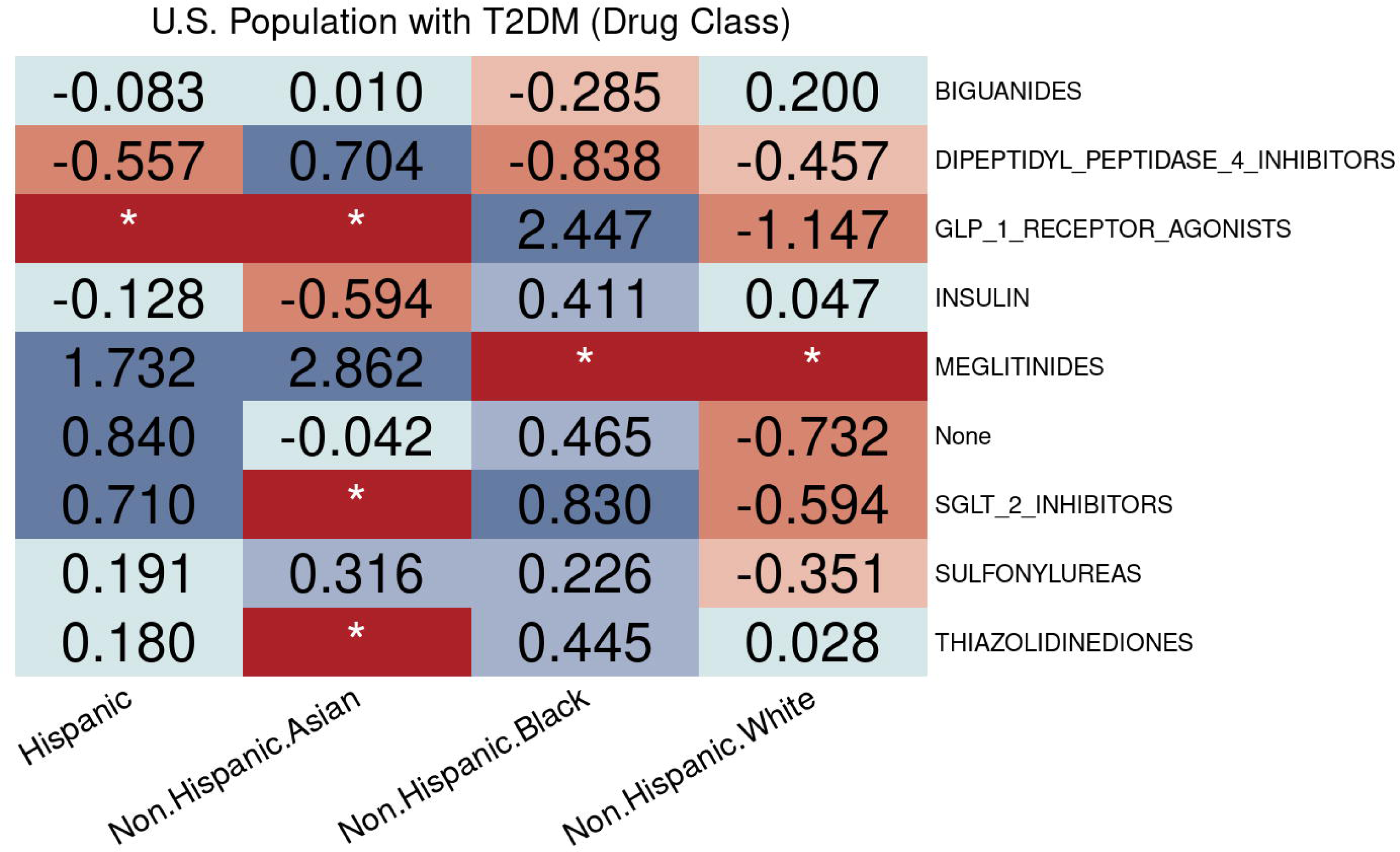
Racial/Ethnic equity evaluation on hyperglycemic medication utilization. Racial/Ethnic disparities of hyperglycemic medication utilization among diabetic population with HbA1c *≥* 6% who have different types of insurance. A: People with non-private insurance. B: People with private insurance.

**Fig 7.**
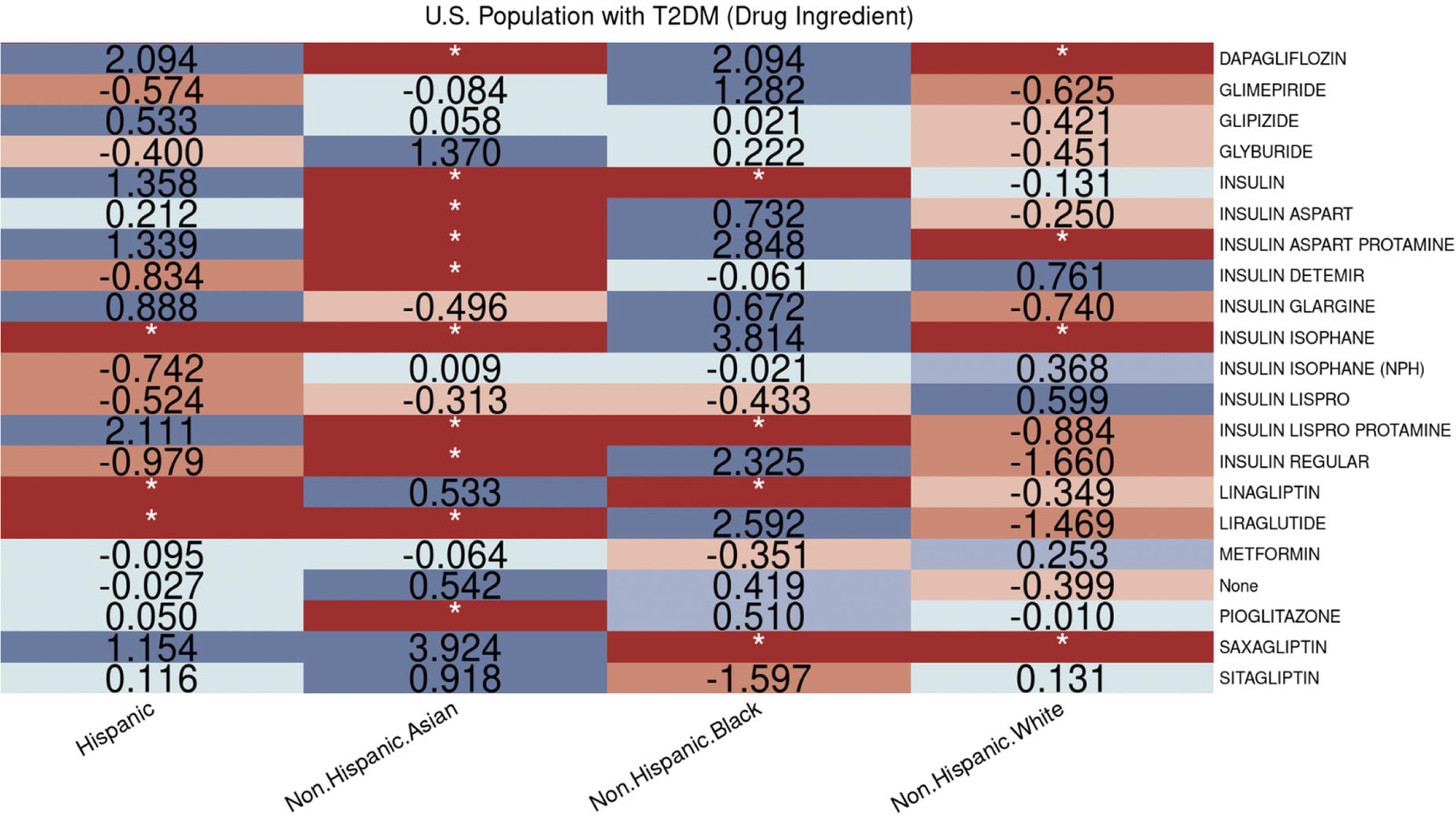
Disparities of ingredient utilization in hyperglycemic medications by race/ethnicity.

We additionally analyzed access and utilization of antidiabetic ingredients, which provides a deeper understanding of the root of disparities and better design interventions to eliminate inequities in healthcare. For example, according to the evaluation of medication classification, it is hard to know whether high-priced insulins were prescribed to non-Hispanic White patients; however, based on evaluation of ingredients, we observed that among different types of long-acting insulins prescribed to non-Hispanic White patients, the more expensive type (i.e., insulin detemir) has a higher rate of being prescribed.

## Discussion

The two approaches evaluate utilization disparity in health care access from two aspects. The logistic regression model is used to compare access and utilization to a reference group while controlling for potential confounders. It can consider the potential effects of drug treatment shifts caused by official guidelines and new knowledge on drugs from new studies, such as cardiovascular outcome trials [31], over time. Medications that entered into the U.S. market earlier may lead to a higher prescribing rate. The equity-metric based visualization approach evaluates whether a subgroup has adequate access and utilization of healthcare with respect to the specified target population. It helps eliminate the bias from unbalanced subgroup proportions in the target population. But the equity approach can only address a limited number of discretized control factors. As the number of factors of interest grows there must be more data to adequately represent the subgroups, and the visualizations become less effective. In general, the logistic regression model focuses on the sameness across people irrespective of being rational or not; our fairness-based visualization approach instead focuses on treating populations fairly but differently.

We observed access and utilization disparities derived from demographic and socioeconomic factors. For instance, both race/ethnicity and insurance status may determine access to some classes of medications, especially the newer ones.

Our results also revealed the importance of multivariate analysis conditioned on disease-specific health conditions such as HbA1c levels of patients to evaluate access to healthcare across subgroups defined over demographic and socioeconomic attributes that should not influence health outcomes. For instance, inadequate access to DDP4-Is in the Hispanic community only existed if they have non-private insurance.

However, some disparities may be associated with disease and treatment-specific factors. For example, the vaccination disparities by age groups in HPV vaccination may be due to the CDC recommendations [32]. CDC does not recommend HPV vaccination for people over age 26 but some people may get it at an older age if not adequately vaccinated when younger. Additionally, whether one receives the third dose of the HPV vaccine depends on the time between the first and second doses. So, collaboration with physicians and endocrinologists is necessary to discover more meaningful clinical-relevant findings.

Our experiment had some limitations. First, prescription medication information was self-reported by participants, which might be subject to reporting bias. Second, some medication/ingredient class samples were not large enough in the subgroup analyses to provide further exploration. Furthermore, since NHANES does not provide all health conditions required to calculate CCI, which is associated with prescribing decisions of antidiabetic drugs, more comprehensive data resources should be examined.

With the use of the semantic infrastructure, it is possible to mitigate some of the limitations, such as further expanding the drug selection criteria, since the NHANES Knowledge Graph expands the original data to include content from existing databases such as ICD10-CM [33] and RxNorm [34]. However, we have not leveraged these expansions in our current analysis since our original goal was to be able to compare the results of analysis generated from datasets manually derived from raw NHANES data against analysis from datasets derived from our semantic infrastructure.

Further research includes the study of access and utilization of other healthcare, such as hospitalization and COVID-19 vaccination, on a more comprehensive list of socioeconomic and demographic characteristics that may influence health status and health outcomes of populations with health care needs. Moreover, a semantic-based evaluation framework that incorporates with our approach should be developed to automatically summarize and explain findings to physicians and policymakers to support public health and clinical decision making and interventions.

## Conclusion

Our findings provide evidence of inequitable accessibility and utilization of hyperglycemic medications and CDC-recommended vaccines, contributed by demographic characteristics (e.g., race and ethnicity), and socioeconomic characteristics (e.g., insurance types and education attainments). However, determinants of access to different healthcare are not the same, requiring disease-level analyses. These discoveries indicate the need and potential interventions to reduce preventable disparities in health care access and utilization among different populations and thus allow every person to live healthier lives.

We developed a powerful equity-analysis methodology and infrastructure that can be generalized to investigate disparities in other types of health care access using NHANES. The proposed equity-focused methodology identifies both potential determinants of access to health service and impacted subgroups. To facilitate this and future public health investigations, our semantic infrastructure supported the data preparation steps of this experiment. In place of combining and normalizing variables from several datasets across the NHANES cycles used in this analysis, this process was facilitated by user interfaces and the NHANES ontology. By visually browsing ontology terms, we could select sets of variables of interest based on their semantic meaning (e.g. “insurance coverage”), instead of manually combining several variables (such as seven variables that characterize insurance coverage). As we expand the NHANES ontology to cover additional NHANES datasets and cycles, we expect the usefulness of this approach to be reused in new studies that use NHANES data.

## Data availability

- **NHANES raw data**: Freely available at https://www.cdc.gov/nchs/nhanes/index.htm
- **Semantic Data Dictionaries and Ontologies:** https://github.com/tetherless-world/nhanes-hadatac

## Data Availability

NHANES raw data are freely available at
https://www.cdc.gov/nchs/nhanes/index.htm. Semantic Data Dictionaries and Ontologies are available in the
https://github.com/tetherless-world/nhanes-hadatac .

## Acknowledgments

This work was partially supported by IBM Research AI Horizons Network.

## Footnotes

1 https://github.com/tetherless-world/nhanes-hadatac

